# Peripheral Venous Pressure Accurately Evaluates Congestion in Constrictive pericarditis

**DOI:** 10.1101/2025.01.31.25321509

**Authors:** Xue Xia, Jianzhong Zhou

## Abstract

**Background:** The primary clinical characteristic of initial constrictive pericarditis(CP) is a cardiac decreased diastolic function, which results in venous congestion, limitation of venous return, and elevated central venous pressure(CVP).

Objective was to evaluate the relationship between peripheral venous pressure (PVP) and CVP with the aim to assess the diagnostic value of PVP in CP.

**Methods:** Subjects underwent invasive haemodynamic and low invasive peripheral venous pressure assessment and also underwent physical examination, biochemical indices to assess venous congestion and echocardiography to assess cardiac structure and function. Patients were divided into 2 groups by CVP > 22 cmH2O(Modal 1 defined as CVP > 22 cmH2O; Modal 2 defined as CVP≦22cmH2O).

**Results:** Of the 36 patients, PVP and CVP had a significant positive association (Pearson’s = 0.831; P < 0.0001), with CVP = 0.962 + 0.907PVP as the regression equation.PVP was negatively correlated with FIB-4 (Pearson’s = −0.425; P=0.01), while it was not statistically significantly correlated with Pro-BNP, albumin, serum Na, hematocrit, hemoglobin, total bilirubin, direct bilirubin, or ARPI. Modal 1 and 2 groups were separated based on CVP > 22 cmH2O. By comparing the area under the curve (AUC) of the participants’ working mass, it was definitively established that 24 cmH2O was the ideal PVP threshold for the diagnosis of CP.

**Conclusions:** In Constrictive Pericarditis, PVP is a reliable, minimally invasive, and accurate technique for calculating CVP, which is remarkably positively correlated with CVP. Measurement of PVP is therefore important in the assessment of congestion and helps in the early diagnosis of Constrictive Pericarditis when PVP is greater than 24 cmH2O.

## INTRODUCTION

Constrictive pericarditis (CP) is often missed or misdiagnosed because of its insidious onset and infrequency. Many people with cerebral palsy do not receive a diagnosis in the early stages of the illness, even though early detection may enhance patients’ quality of life. High venous pressure congestion is a feature that can manifest early in constrictive pericarditis and is often accompanied by elevated CVP. Whereas right heart catheterization is costly, intrusive, time-consuming, and not easily accessible at the patient’s bedside, better bedside venous pressure assessment tools have become a clinical necessity. Clinical assessment can usually be done by physical examination, biochemical markers, and imaging, including peripheral edema and jugular vein rages on physical examination, biochemical tests (e.g.natriuretic peptide, the Liver Fibrosis Markers, and Scores), and imaging (e.g., CT, Echocardiography) **^Error! Reference source not found.Error! Reference source not found.^**[2][3]**^Error! Reference source not found.^**^etc^. An invasive hemodynamic assessment was performed when the above could not be determined. PVP and CVP have been shown to be substantially positively associated in numerous studies of research; therefore, PVP can be used as a marker of CVP as a substitute for CVP for bedside assessment of volume condition in patients with constrictive pericarditis and diagnosis of high venous pressure congestion.

The accuracy of the current physical examination for assessing high venous pressure congestion is limited, and researchers have shown a salient relationship between CVP and PVP in heart failure, Fontan circulation[5][6]**^Error! Reference source not found.Error! Reference source not found.^**, and current investigations[9]have revealed that the assessment of congestion by PVP is more accurate than conventional congestion assessment methods. Therefore, this study aimed to assess the state of venous congestion caused by diastolic restriction due to diseased pericardium through the application of PVP in constrictive pericarditis patients and analyzing the capacity of PVP to assess CVP to improve the identification of patients with constrictive pericarditis as well as the guidance of volume management.

## METHODS

### Study Population

This was a single-centre, retrospective study comprising 36 patients with constrictive pericarditis who were diagnosed definitively by pathology after pericardiectomy at the First Affiliated Hospital of Chongqing Medical University (the CP group) from January 2012 to June 2024 and before pericardiectomy, we simultaneously performed routine congestion assessment, CVP and PVP measurements and assessed PVP’s ability to detect CVP; we also measured PVP values in 26 patients with Non-Constrictive Pericarditis served as controls (the control group) and examined if the PVP of the CP and control groups differed from one another. People with active tumors, active hepatitis or viral cirrhosis, severe myocardial ischemia, venous thrombosis, phlebitis, severe renal failure (creatinine < 3 mg/dl or on hemodialysis), malnourishment, or known restrictive cardiomyopathy were not included in the previous study protocol[10].To ensure consistency of results, at least two of the three full-time, experienced researchers who conducted the physical examinations for this study examined each patient; all reviewers were not provided with information on CVP, PVP, and related examination tests. All study procedures complied with the ethical principles of the Declaration of Helsinki, and all patients provided written informed consent. The Institutional Review Board of the First Hospital of Chongqing Medical University approved the study protocol.

### Measurement of PVP

During the study, to prevent further punctures just for this research, we mainly gauged PVP through the 22 G peripheral venous infusion line situated as the venous access in the upper limb. In this study, the manual measurement method was used, and the measurement method was that the patient took the lying position during measurement. The position was zeroed before measurement (right atrium level, i.e., the intersection of the axillary midline and the fourth intercostal space). Meanwhile, the upper limbs were abducted by 45°. The peripheral venous pipeline was connected to saline, and after the intravenous drip of the liquid was stabilized, the infusion set was separated from the bottle, and the drop of the water column was observed. The vertical distance was determined as the PVP.

### Measurement of CVP

After the measurement of PVP, the patients in this study entered the operating room for central venous catheterization. All patients were placed in the lying position, the right internal jugular vein was selected as the puncture point, and a central venous catheter was placed by a puncture after local anesthesia with a disposable central venous puncture kit, the CVP port was situated in the superior vena cava, or right atrium and a pressure transducer was connected to measure the CVP value at the end of the expiration.

### Statistical Analysis

Categorical variables are expressed as n (%), and continuous variables are expressed as mean ± standard deviation or median (inter-quartile ranges). This research was conducted to investigate the extent of PVP and CVP’s association, with Pearson’s correlation coefficients assessing the correlation and The Intraclass Correlation Coefficient (ICC) assessing the consistency of the data. The working characteristic curves of subjects with PVP were plotted by grouping them with a cut-off value of CVP ≥22 cmH2O[12], and the differences in PVP between CP and non-CP patients were analyzed by t-test. The working characteristic curves of subjects with PVP were plotted by grouping them with a cut-off value of CVP ≥22 cmH2O, and the differences in PVP between CP and non-CP patients were analyzed by t-test. P values <0.05 were regarded as significant, and all statistical tests were two-sided. GraphPad Prism 10.1.2 (GraphPad Software, Inc., La Jolla, CA, USA) and SPSS 27.0.0.0 (190) (IBM, Armonk, NY, USA) were utilized for statistical analysis.

## RESULTS

### Baseline Characteristics

We analyzed data from 36 patients, and all patients had been diagnosed with CP and underwent pericardiectomy. Table 1 displays the clinical features of the patients. The median time to diagnosis for these 36 patients was 3 months (2,12). The median age was 59 years,32 patients were male (63%),11% had an ischemic pathogenesis of heart failure, 25% had a history of Tuberculosis, and 86% had the New York Heart Association functional Class ≥III. In our cohort,78% exhibited dyspnea, and 44% showed chest tightness. Additionally,33% had peripheral edema, and 56% had Jugular venous congestion on physical examination. Patients with symptomatic signs such as peripheral edema, dyspnoea, chest tightness, and jugular vein rages had higher PVP and CVP, but there was no statistically significant difference.

**Table 1.**
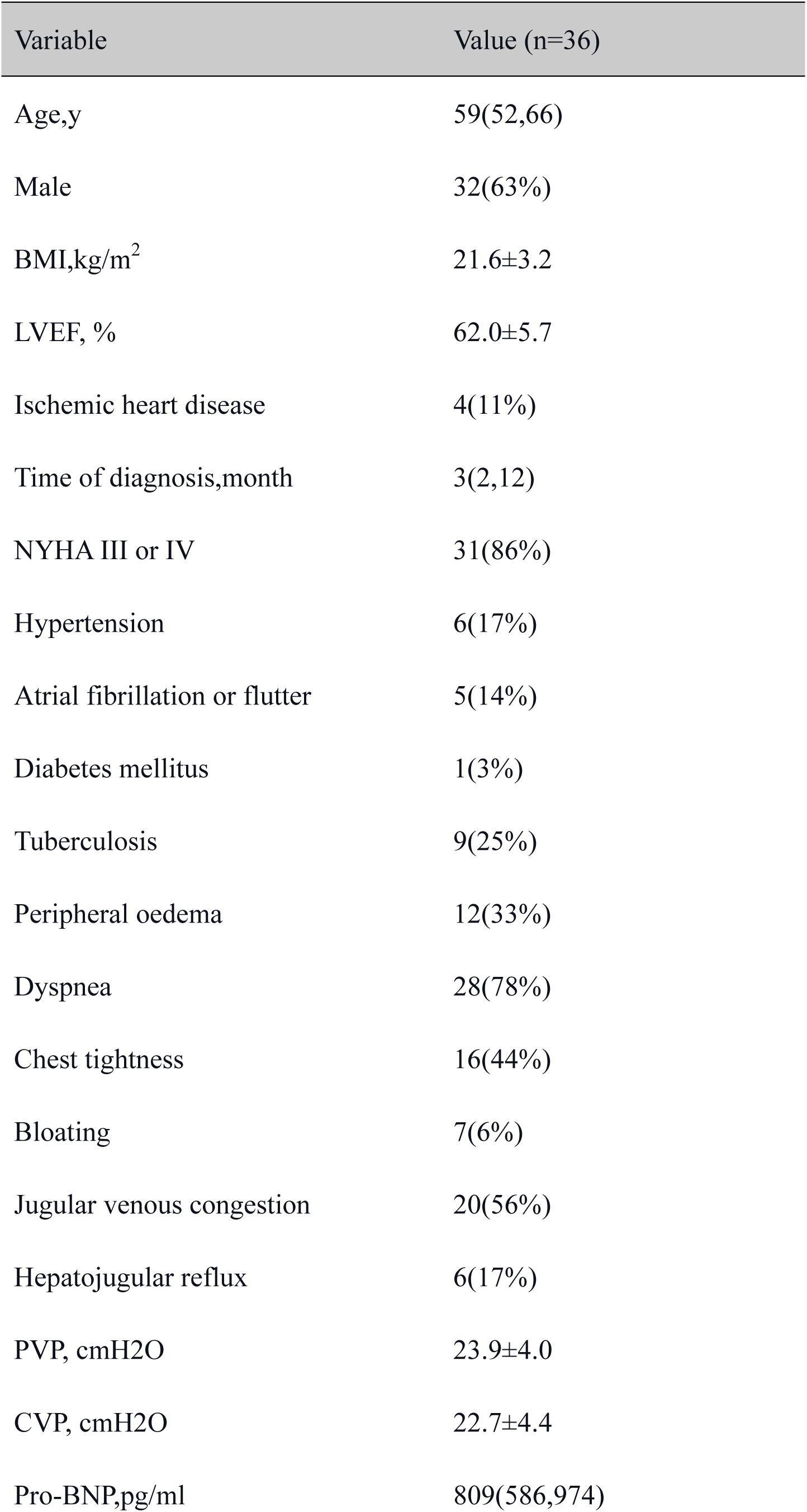
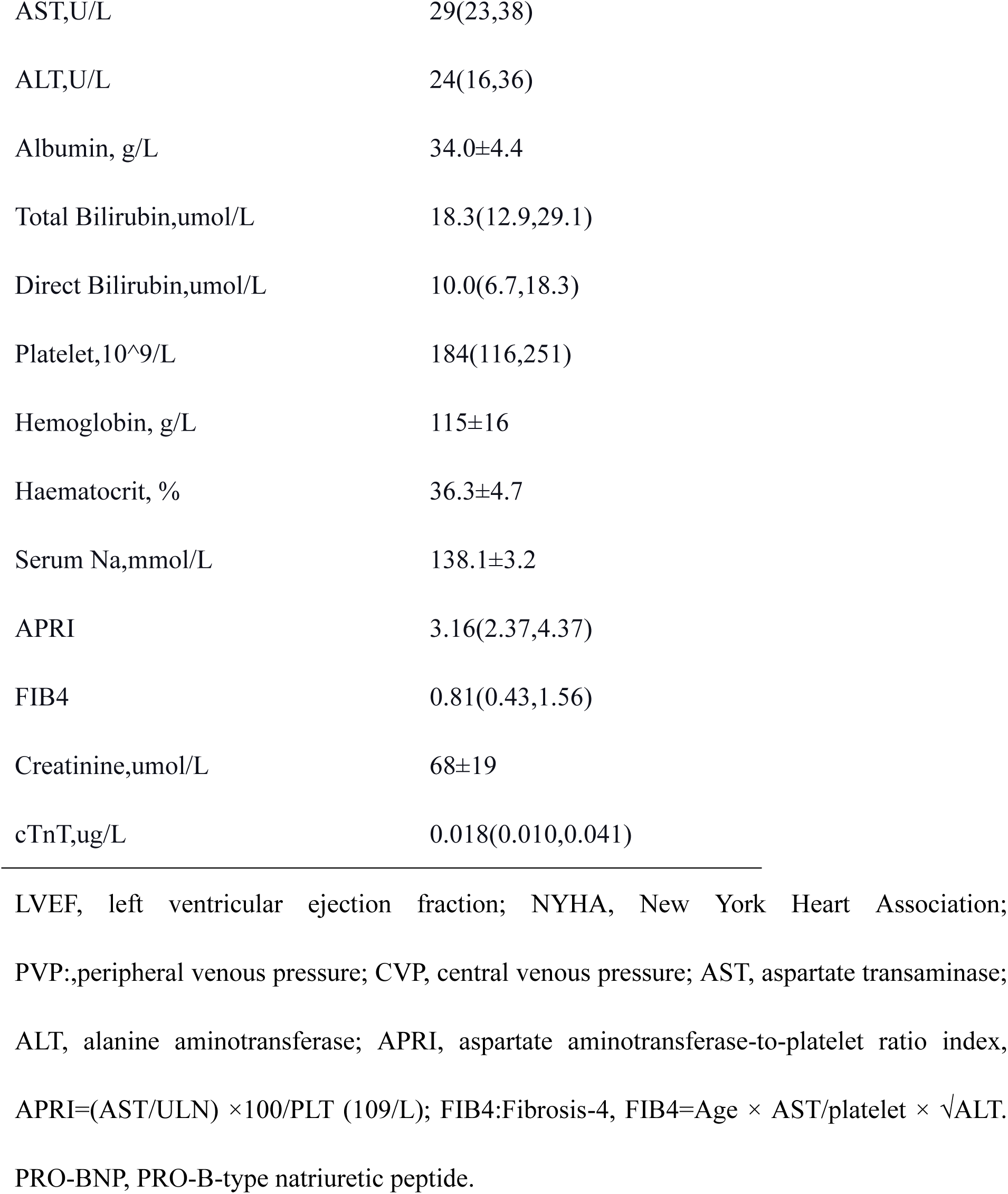
Baseline Characteristics of CP.

### Comparison of conventional assessments and peripheral venous pressure

The mean CVP was 22.7±4.4 cmH2O, and the mean average PVP was 23.9±4.0 cmH2O, (Figure 1A). CVP and PVP are strictly linked (Figure 1B; r=0.831, P<0.0001). The equation CVP = 0.962+0.907* PVP describes a best-fit linear correlation. The Intraclass Correlation Coefficient (ICC) was 0.886(P<0.001) for CVP and PVP, which indicated the CVP was strongly consistent with PVP. Comparing conventional assessment of congestion with PVP and CVP, PVP and CVP were not significantly correlated with Pro-BNP, Albumin, Serum Na, Haematocrit, Hemoglobin, Total Bilirubin, Direct Bilirubin, and ARPI, and they were both negatively correlated with FIB-4 (Figure 2 CVP r=-0.473, P=0.0035; PVP r= −0.425, P=0.01).

**Figure 1.**
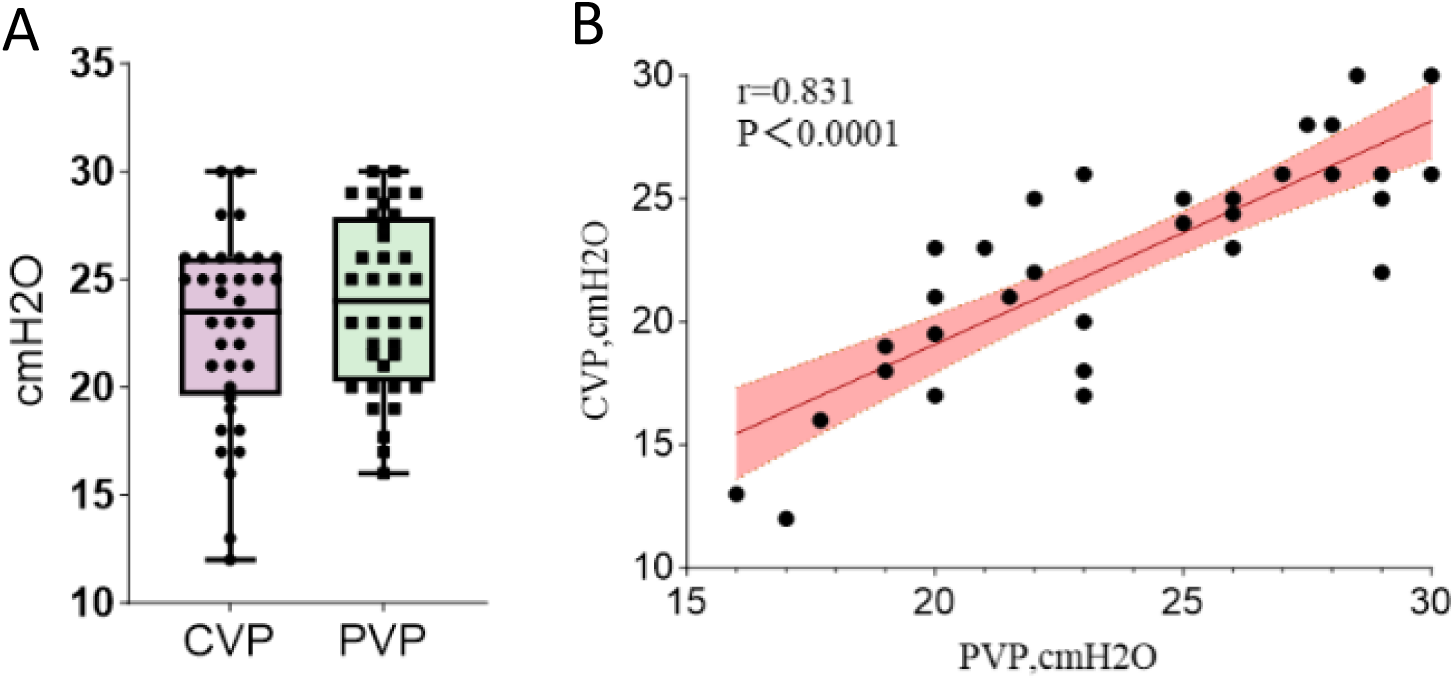
Correlation of PVP with CVP. (A) Comparing PVP with CVP(23.9±4.0 cmH2O vs 22.7±4.4 cmH2O) in CP. (B) Scatter plot demonstrating the correlation between peripheral venous pressure (PVP) and central venous pressure (CVP). Pearson’s r=0.831, P<0.0001.

**Figure 2.**
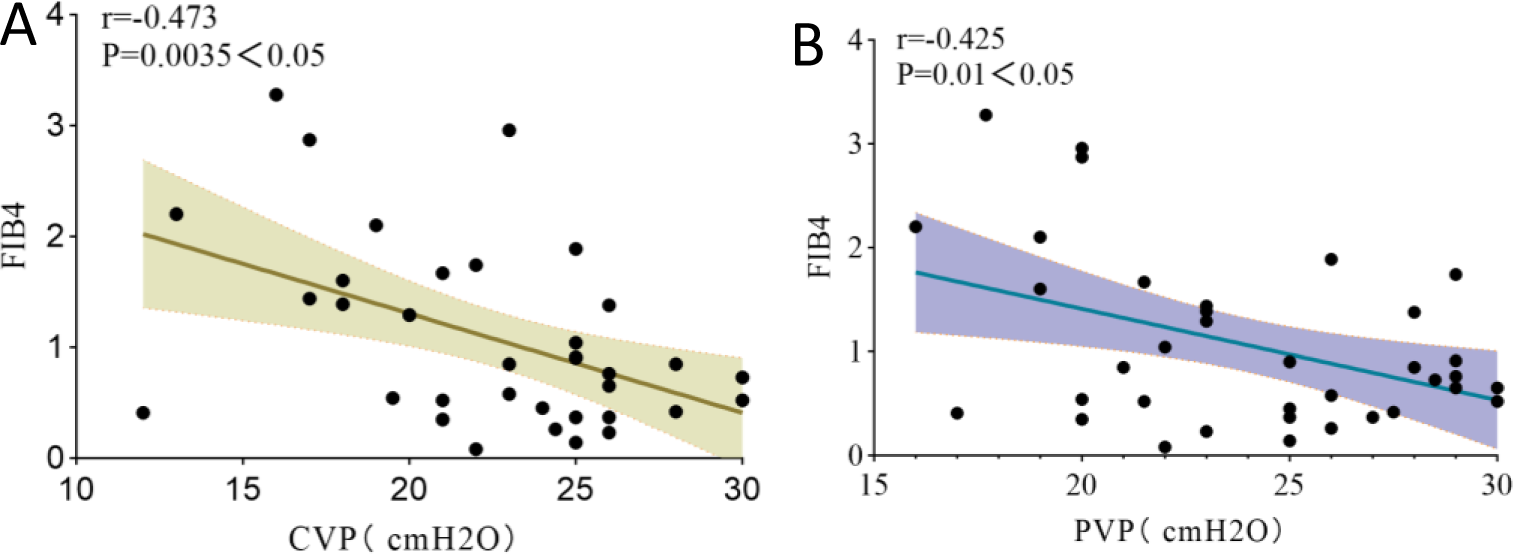
Relationship between FIB4 in PVP and CVP. (A)Scatter plot demonstrating the correlation between Fibrinogen 4 (FIB4) and central venous pressure (CVP), pearson’s r=-0.473, P<0.05. (B)Scatter plot demonstrating the correlation between Fibrinogen 4 (FIB4) and peripheral venous pressure (PVP). pearson’s r=-0.425, P<0.05.

PVP for the control group was 12.5±5.5 cmH2O, which was significantly lower than the CP’s (Figure 3; 12.5±5.5vs23.9±4.0, P<0.0001). In addition, divided into two groups with CVP; 22 cm H2O (Table 2)[12], patients in the high CVP group(Modal 1) were more likely to have peripheral edema (10 vs 2, P = 0.03) and had a higher PVP and lower FIB-4 (Figure 4A,26 ± 3 vs 21 ± 3, P < 0.001; Figure 4B,0.65(0.40,0.91) vs 1.4(0.5,2.1), P = 0.02). The time to diagnosis was greater in model 2 than in model 1, despite the fact that there was no statistically significant difference between the two models (3vs6, P=0.48). According to the AUC (Figure 5), a PVP cut-off value of 24 cmH2O had a sensitivity of 81 % and a specificity of 93 % for diagnosing patients with constrictive pericarditis. This corresponds to an accuracy of 86% in detecting constrictive pericarditis, with a negative predictive value of 78% and a positive predictive value of 94%. The receiver operating characteristic curves are shown in Figure 8, showing the sensitivity and specificity for different PVP values with an area under the curve of 0.8857(Figure 5).

**Figure 3.**
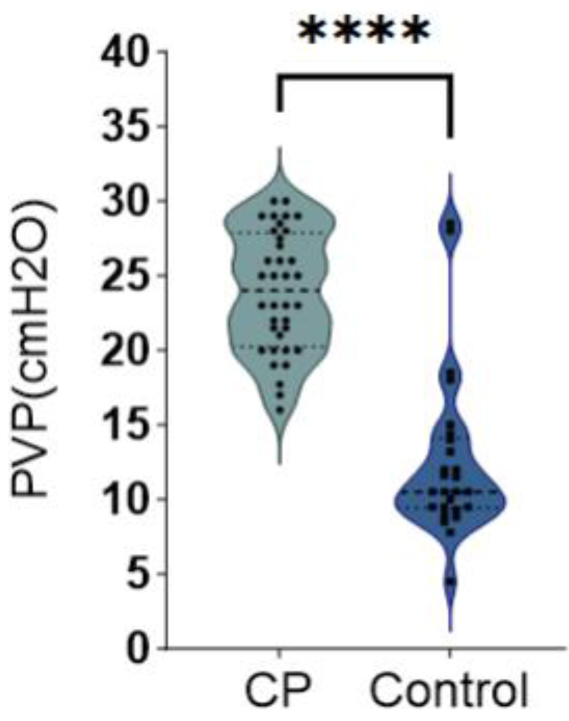
Comparison of PVP between the experimental group and the control group showed a statistically significant difference. P<0.001, 95%CI(9.0116,13.8529).

**Figure 4.**
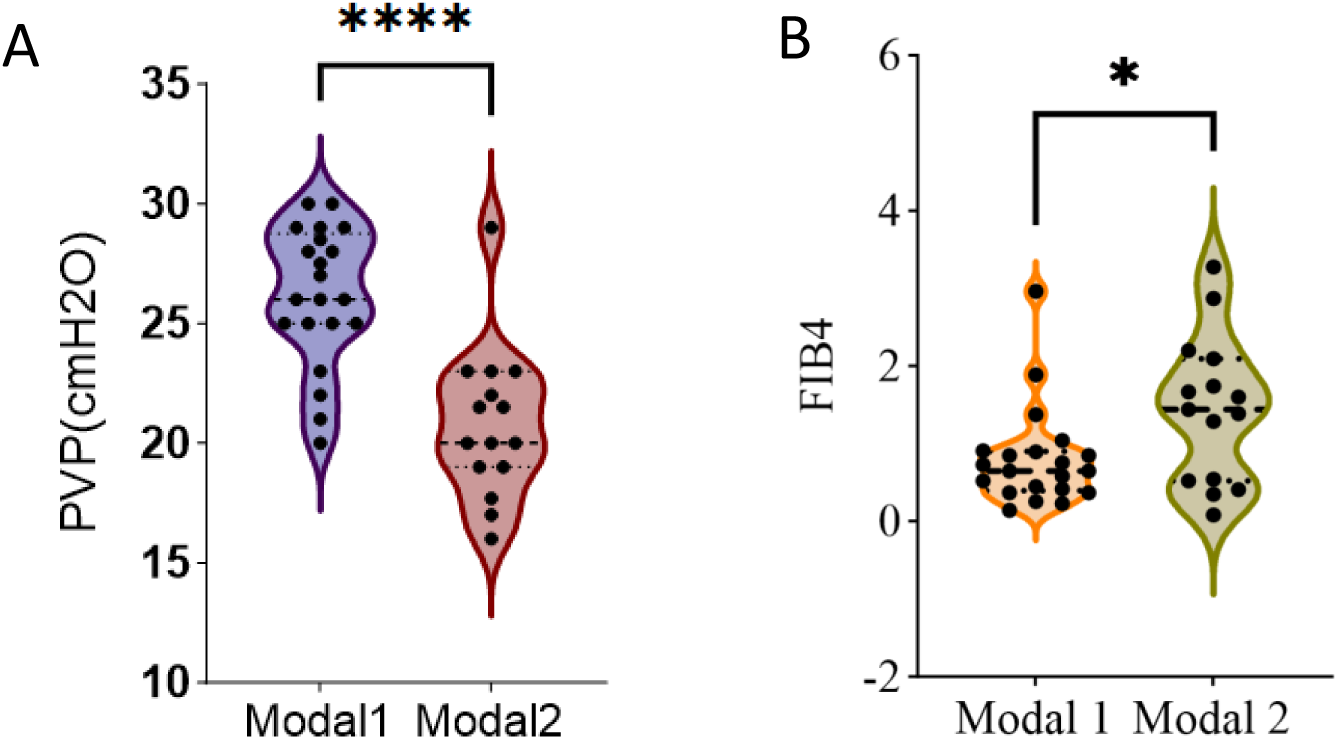
Comparison of groups with CVP > 22 cmH2O. Modal 1:CVP; 22 cm H2O; Modal 2:CVP≤22 cm H2O. (A)Comparison of PVP between the Modal 1 and the Modal 2 demonstrated a statistically significant difference, P < 0.001, 95%CI(3.3514,7.4695). (B)Comparison of FIB4 between the Modal 1 and the Modal 2 displayed a minor linkage, P=0.02<0.05, 95%CI(−1.16,−0.009).

**Figure 5.**
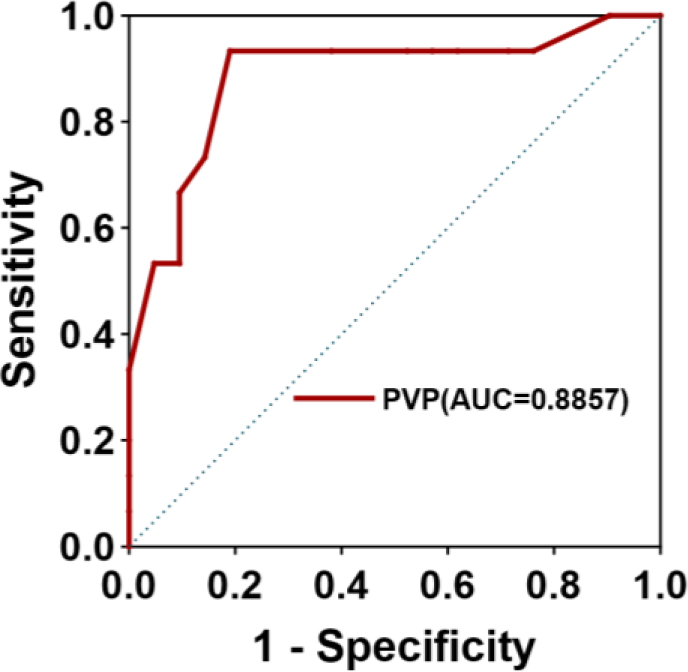
Receiver Operating Characteristic Curve for PVP measurement as a tool for detecting elevated CVP in the CP (CVP > 22 cm H2O).

**Table 2.**
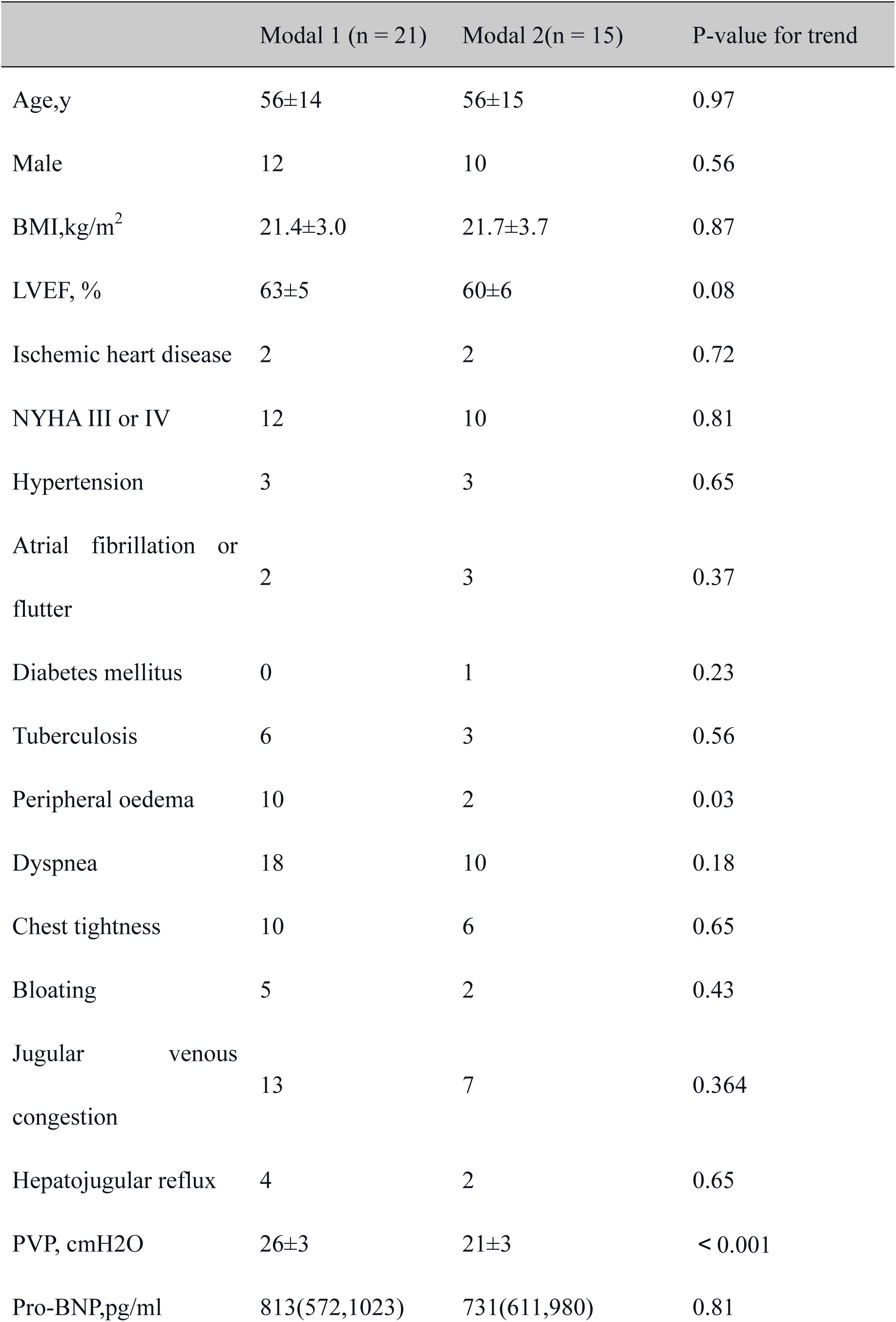
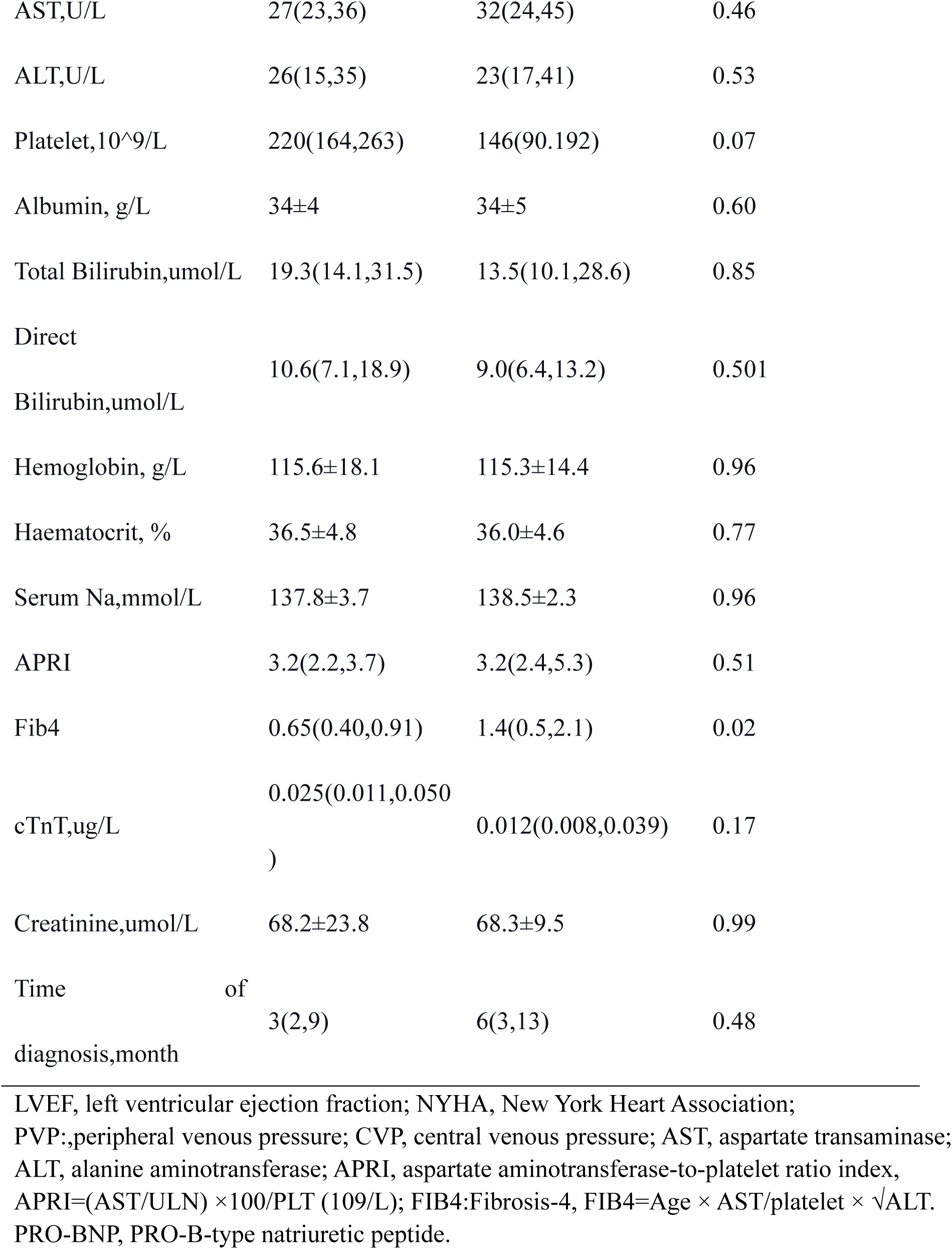
Divided into two groups with CVP>22 cm H2O.

## DISCUSSION

This is a single-center, retrospective study to assess the connection between peripheral and central venous pressure to support the diagnosis of venous congestion in patients with CP and to improve its diagnostic accuracy. This research found that in patients with constrictive pericarditis, central venous pressures examined invasively were similar to peripheral venous pressures measured from the upper limbs. These findings support the diagnosis and treatment of constrictive pericarditis and show that non-invasive evaluation of the condition can be performed in emergency and outpatient settings. This study demonstrated that PVP is highly correlated and consistent with CVP in constrictive pericarditis. Previous studies have shown a logical correlation between CVP and PVP in other patient populations, including gastrointestinal surgery**Error! Reference source not found.**, neurosurgery**Error! Reference source not found.**, Fontan circulation**^Error! Reference source not found.^**, and acute heart failure[5]showing excellent correlation, and that the ability of PVP to assess CVP is superior to other routine assessments.

In addition, most previous studies measured peripheral venous pressure using a pressure monitoring transducer connected to a monitor, which is expensive and challenging to implement in general wards and outpatient clinics. In contrast, our research mainly used manual measurement of peripheral venous pressure, which is easier to implement in clinical practice and avoids the risk of infection, embolism, and vascular injury. It is also more straightforward, more economical, and more widely applicable. According to the CVP grouping, the AUC of the PVP cut-off value was 88.57%, with high diagnostic efficacy. In the future, clinics can consider patients suffering from peripheral edema, Dyspnea, Jugular venous congestion, and other symptoms and signs. If echocardiography and CT suggest insufficient evidence of CP, the PVP value can be measured, and if it is higher than 24 cmH2O, the diagnosis of CP can be made.

Elevated venous pressure in HF, primarily HFpEF, results in passive hepatic stasis, which in consequence produces perisinusoidal edema, hepatocellular atrophy, hepatic stasis, and a subsequent increase in liver stiffness. Fibrinogen 4 (FIB4) score is a non-invasive marker for assessing non-alcoholic fatty liver disease (NAFLD). It is determined by biochemical values, including platelets, alanine aminotransferase (ALT), aspartate transaminase (AST), and age (FIB4=Age × AST/platelet × √ALT).BIELECKA and SHIRAKABE[15]**^Error! Reference source not found.^**showed that elevated FIB4 was associated with cardiac function, renal impairment, and fluid overload; Mitsutaka[16]noted that elevated FIB4 was connected with right ventricular dysfunction in HFpEF and increased risk of future MACE; Yamada**^Error! Reference source not found.^** also pointed out that FIB4 was an AHF survival independent predictor and that FIB4 had a high predictive value for HFpEF, HFmrEF and CAD death. In previous research, it was also explained that advanced fibrosis with FIB-4 score is a risk factor for cardiovascular disease mortality in patients with NAFLD[18][19].In our study, FIB4 was negatively correlated with both PVP and CVP. Still, in combination with the duration of illness in the CP group, when the longer the time required for the diagnosis of CP, the longer the heart is subjected to diastolic limitation due to pericardial constriction, the longer the liver is bruised, and then the stiffer the liver subsequently becomes. The FIB4 value assessed is even higher. The higher PVP and lower FIB4 in those with a shorter time to diagnosis also reflect the fact that the venous congestion symptoms of CP can occur earlier by PVP before it involves the liver and causes pathology. This study also suggests that PVP may be more effective for the early diagnosis of CP, but a large amount of clinical data is also needed for future validation assessment.

This study demonstrates that PVP is a reliable alternative for estimating CVP in CP when non-invasive estimation of venous congestion is difficult by physical examination and echocardiographic and CT assessment. Healthcare providers may be better able to identify this population using reliable and accurate CVP measurement techniques, and this may have the potential to benefit perioperative blood volume management in patients with CP.

### Limitations

Even though our data indicates that PVP and CVP are significantly correlated with constrictive pericarditis, there are several limitations to consider. Firstly, the study was a single-center study with a small sample population, and its replicability in a larger multicentre group of patients with constrictive pericarditis has yet to be validated. Secondly, this research only paired measures of PVP and CVP at one time period rather than examining this correlation at several time points during hospitalization. Thirdly, the PVP cut-off values derived in this study were not validated in an independent cohort. Fourthly, for physical examination, measurements that are biased will affect the accuracy of routine assessment, and we did not conduct a comprehensive echocardiographic analysis, such as tissue Doppler index and hepatic blood flow patterns[20]. Ultimately, concerning the measurement of each parameter, the impact of additional unmeasured confounding factors cannot be completely ruled out.

## CONCLUSIONS

PVP is a simple, cost-effective, minimally invasive, and reliable method of estimating CVP in Constrictive Pericarditis, with a marked positive correlation between PVP and CVP and higher PVP in the CP group; therefore, early PVP assessment has clinical value in supporting the diagnosis of Constrictive pericarditis. Utilizing this PVP in clinical settings could enhance the way congestion is currently assessed and assist in diagnosing constrictive pericarditis and managing volume guidance during the perioperative period.

## Data Availability

the data that support the findings of this study are available from the corresponding author upon reasonable request.

## Acknowledgments

This research was supported by the Department of Cardiovascular Medicine and Department of Cardiothoracic Surgery of the First Affiliated Hospital of Chongqing Medical University.

## Supplemental Material

Simplified PVP measurement Graphical Abstract (By Figdraw)

## Nonstandard Abbreviations and Acronyms

CP: constrictive pericarditis central venous pressure

CVP PVP: peripheral venous pressure

FIB-4: Fibrosis-4

Pro-BNP: PRO-B-type natriuretic peptide

ARPI: Aspartate aminotransferase-to-platelet ratio index

ICC: Intraclass Correlation Coefficient

HF: Heart Failure

HFpEF: Heart Failure with preserved ejection fraction

HFmrEF: Heart Failure with mid-range ejection fraction

LVEF: left ventricular ejection fraction

NYHA: New York Heart Association

NAFLD: Non-alcoholic fatty liver disease

ALT: Alanine aminotransferase

AST: Aspartate transaminase

MACE: Major Adverse Cardiovascular Events

CAD: Coronary Artery Disease

BMI: Body mass index

CT: Computed Tomography

AUC: Area Under the Curve

## REFERENCES

[1] Mullens W, Damman K, Harjola VP, Mebazaa A, Brunner-La Rocca HP, Martens P, Testani JM, Tang WHW, Orso F, Rossignol P, et al. The use of diuretics in heart failure with congestion - a position statement from the Heart Failure Association of the European Society of Cardiology. Eur J Heart Fail. 2019;21(2):137–155. doi: 10.1002/ejhf.1369.

[2] Adler Y, Charron P, Imazio M, Badano L, Barón-Esquivias G, Bogaert J, Brucato A, Gueret P, Klingel K, Lionis C, et al. ESC Scientific Document Group. 2015 ESC Guidelines for the diagnosis and management of pericardial diseases: The Task Force for the Diagnosis and Management of Pericardial Diseases of the European Society of Cardiology (ESC)Endorsed by: The European Association for Cardio-Thoracic Surgery (EACTS). Eur Heart J. 2015;36(42):2921–2964. doi: 10.1093/eurheartj/ehv318.

[3] Welch, TD. Constrictive pericarditis: diagnosis, management and clinical outcomes. HEART. 2018; 104 (9): 725–731. doi: 10.1136/heartjnl-2017-311683.

[4] Tseng CH, Huang WM, Yu WC, Cheng HM, Chang HC, Hsu PF, Chiang CE, Chen CH, Sung SH. The fibrosis-4 score is associated with long-term mortality in different phenotypes of acute heart failure. EUR J CLIN INVEST. 2022; 52 (12): e13856. doi: 10.1111/eci.13856.

[5] Bielecka-Dabrowa A, Sakowicz A, Banach M, Slot M. An Elevated FIB-4 Score is associated with the severity of heart failure EUR HEART J. 2024; 45 (Supple1): doi: 10.1093/eurheartj/ehae666.1168.

[6] Nagao K, Maruichi-Kawakami S, Aida K, Matsuto K, Imamoto K, Yukawa H, Kanazawa T, Kobayashi Y, Takahashi N, Ito H, et al. Association Between the Liver Fibrosis Markers and Scores, and Hemodynamic Congestion Assessed by Peripheral Venous Pressure in Patients With Acute Heart Failure. J Am Heart Assoc. 2023; 12 (21): e030788. doi: 10.1161/JAHA.123.030788.

[7] Masutani S, Kurishima C, Yana A, Kuwata S, Iwamoto Y, Saiki H, Ishido H, Senzaki H. Assessment of central venous physiology of Fontan circulation using peripheral venous pressure. J THORAC CARDIOV SUR. 2017; 153 (4): 912–920. doi: 10.1016/j.jtcvs.2016.11.061.

[8] Tan W, Small A, Gallotti R, Moore J, Aboulhosn J. Peripheral venous pressure accurately predicts central venous pressure in the adult Fontan circulation. INT J CARDIOL. 2021; 326 77–80. doi: 10.1016/j.ijcard.2020.11.007.

[9] Wengrofsky A, Flint S, Vlismas P, Wiesenfeld E, Rochlani Y, Chavez P, Vukelic S, Murthy S, Madan S, Saeed O, et al. PERIPHERAL VENOUS PRESSURE MEASURED WITH A NOVEL MINIATURE PRESSURE TRANSDUCER TO PREDICT CHRONIC HEART FAILURE READMISSION J AM COLL CARDIOL. 2024; 83 (13): 308. doi: 10.1016/s0735-1097(24)02298-8.

[10] Maruichi-Kawakami S, Nagao K, Aida K, Matsuto K, Imamoto K, Tamura A, Takazaki T, Nakatsu T, Tanaka M, Nakayama S, et al. Peripheral Venous Pressure Measurements to Evaluate Congestion in Heart Failure. J CARD FAIL. 2023; 29 (9): 1319–1323. doi: 10.1016/j.cardfail.2021.11.018.

[11] Raju S, Crim W, Buck W. Factors influencing peripheral venous pressure in an experimental model. J VASC SURG-VENOUS L. 2017; 5 (6): 864–874. doi: 10.1016/j.jvsv.2017.05.024.

[12] Konik E, Geske J, Edwards W, Gersh B. Pericardiectomy as a diagnostic and therapeutic procedure. BMJ Case Rep. 2016 10.1136/bcr-2016-217563

[13] Kim SH, Park SY, Cui J, Lee JH, Cho SH, Chae WS, Jin HC, Hwang KH. Peripheral venous pressure as an alternative to central venous pressure in patients undergoing laparoscopic colorectal surgery. BRIT J ANAESTH. 2011; 106 (3): 305–11. doi: 10.1093/bja/aeq399.

[14] Bombardieri AM, Beckman J, Shaw P, Girardi FP, Ma Y, Memtsoudis SG. Comparative utility of centrally versus peripherally transduced venous pressure monitoring in the perioperative period in spine surgery patients. J Clin Anesth. 2012;24(7):542–8. doi: 10.1016/j.jclinane.2012.03.005.

[15] Shibata N, Sumi T, Umemoto N, Kajiura H, Inoue S, Iio Y, Sugiura T, Taniguchi T, Asai T, Yamada M, et al. P5410Combination assessment of renal and hepatic dysfunction improves the predictability of prognosis in patients with acute decompensated heart failure. EUR HEART J. 2019; 40 (Supple1): doi: 10.1093/eurheartj/ehz746.0368.

[16] Shirakabe A, Okazaki H, Matsushita M, Shibata Y, Shigihara S, Nishigoori S, Sawatani T, Tani K, Kiuchi K, Otsuka Y, et al. Clinical Significance of the Fibrosis-4 Index in Patients with Acute Heart Failure Requiring Intensive Care. Int Heart J. 2021;62(4):858–865. doi: 10.1536/ihj.20-793.

[17] Nakashima M, Sakuragi S, Miyoshi T, Takayama S, Kawaguchi T, Kodera N, Akai H, Koide Y, Otsuka H, Wada T, et al. Fibrosis-4 index reflects right ventricular function and prognosis in heart failure with preserved ejection fraction. ESC Heart Fail. 2021;8(3):2240–2247. doi: 10.1002/ehf2.13317.

[18] Yamada T, Morita T, Furukawa Y, Tamaki S, Kawasaki M, Kikuchi A, Kawai T, Seo M, Nakamura J, Abe M, et al. P787Long-term prognostic value of the combination of fibrosis-4 index and acute kidney injury in patients with admitted for acute decompensated heart failure EUR HEART J. 2019; 40 (Supple1): doi: 10.1093/eurheartj/ehz747.0386.

[19] Chew NWS, Ng CH, Chan KE, Chee D, Syn N, Tamaki N, Muthiah M, Noureddin M. FIB-4 Predicts MACE and Cardiovascular Mortality in Patients With Nonalcoholic Fatty Liver Disease. Can J Cardiol. 2022;38(11):1779–1780. doi: 10.1016/j.cjca.2022.07.016.

[20] Henson JB, Simon TG, Kaplan A, Osganian S, Masia R, Corey KE. Advanced fibrosis is associated with incident cardiovascular disease in patients with non-alcoholic fatty liver disease. ALIMENT PHARM THERAP. 2020; 51 (7): 728–736. doi: 10.1111/apt.15660.

[21] Pellicori P, Shah P, Cuthbert J, Urbinati A, Zhang J, Kallvikbacka-Bennett A, Clark AL, Cleland JGF. Prevalence, pattern and clinical relevance of ultrasound indices of congestion in outpatients with heart failure. Eur J Heart Fail. 2019;21(7):904–916. doi: 10.1002/ejhf.1383.

